# MRI-based surrogates of brain clearance in narcolepsy type 1

**DOI:** 10.1101/2024.11.04.24316690

**Authors:** Eva M. van Heese, Jari K. Gool, Gert Jan Lammers, Ysbrand D. van der Werf, Matthias J.P. van Osch, Rolf Fronczek, Lydiane Hirschler

## Abstract

Brain clearance involves the drainage of waste molecules from the brain, a process that is suggested to be amplified during sleep. Recently proposed MRI-based methods attempt to approximate human brain clearance with surrogate measures. The current study aimed to explore whether two brain clearance surrogates are altered in narcolepsy. We processed diffusion-weighted and functional resting-state images to extract two surrogates: Diffusion Tensor Imaging Along the Perivascular Space (DTI-ALPS index), and dBOLD-CSF coupling. Both measures were analysed in 12 drug-free, awake people with narcolepsy type 1 and 11 age- and sex-matched controls, as well as in relation to clinical features. We also assessed the correlation between the DTI-ALPS index and dBOLD-CSF coupling. The DTI-ALPS index and dBOLD-CSF coupling amplitude identified were similar to previous research and did not show significant differences between narcolepsy and controls, nor significant relations with severity of excessive daytime sleepiness. We found a significant correlation between the DTI-ALPS index and dBOLD-CSF coupling amplitude. The hypothesis of altered brain clearance in narcolepsy type 1 is not supported by evidence from the current study. The two surrogates correlated with each other, suggesting that both offer different perspectives from the same underlying physiology. Yet, the suitability of the surrogates as brain clearance markers remains debatable. Whereas DTI is not exclusively sensitive to perivascular fluid, dBOLD-CSF coupling is reflecting large-scale CSF motion. Future work should explore other surrogate markers, preferably during sleep, to better understand the possible role of altered brain clearance in narcolepsy type 1 symptomatology.

## Introduction

### Narcolepsy

Narcolepsy type 1 is a primary neurological sleep disorder characterised by excessive daytime sleepiness (EDS), disturbed nocturnal sleep, and cataplexy (i.e. involuntary loss of muscle tone induced by strong emotions). As people suffering from narcolepsy experience strongly reduced daytime functioning and quality of life, it poses a significant burden for patients, partners, and society (Jennum et al., 2012). The underlying pathophysiology of narcolepsy type 1 is deficiency of hypocretin-1, a neuropeptide involved in sleep-wake regulation that is normally mainly produced in the lateral hypothalamus (Nishino et al., 2000; Thannickal et al., 2000). From the hypothalamus, hypocretin-producing cells have widespread projections to the whole brain, with dense connections to the brainstem nuclei and basal forebrain (Ebrahim et al., 2002).

### Brain clearance

Research on brain clearance has recently gained popularity in the context of healthy sleep and sleep disorders. One of the important elements of the brain clearance system involves cerebrospinal fluid (CSF) movement along channels surrounding the blood vessels: the perivascular spaces. These channels are proposed to be the main pathways for CSF-mediated clearance. Brain clearance is suggested to be amplified during sleep based on observations of faster amyloid-β clearance and interstitial fluid (ISF) compartment enlargements in sleeping mice, and larger CSF compartment volume in sleeping humans (Demiral et al., 2019; Xie et al., 2013). Moreover, sleep deprivation in humans has been shown to slow down brain clearance (studied using tracer enrichment after intrathecal injections; (Eide et al., 2021). Currently, different models exist to describe the anatomy and direction of fluid movement underlying brain clearance (Bakker et al., 2019; Carare et al., 2008; Iliff et al., 2012; Zhao et al., 2022).

### Altered brain clearance in narcolepsy?

There are several lines of evidence suggesting the presence of brain clearance alterations in narcolepsy. First, the hypothesis of altered clearance is supported by narcolepsy being characterised by a highly disrupted sleep-wake cycle (i.e. high number of awakenings, altered sleep stage transitions, more time spent in stage 1 non-rapid eye-movement [NREM] sleep, and fragmented NREM sleep; (Pizza et al., 2015; Roth et al., 2013). Brain clearance is suggested to be amplified during NREM sleep, presumably stage 2 and/or 3. We hypothesise that the disrupted sleep-wake cycle in narcolepsy could be related to altered brain clearance.

Second, abnormal levels of CSF biomarkers support the idea of altered clearance in narcolepsy. Spinal CSF measurements are often performed in people with narcolepsy, to confirm hypocretin-1 deficiency. Several studies have used this opportunity to investigate other CSF biomarkers, mostly neurodegenerative markers such as amyloid-β, phosphorylated and total tau, and neurofilament light. A summary of this work is presented in Table 1 (Baiardi et al., 2020; Heier et al., 2014; Jennum et al., 2017; Kallweit et al., 2012; Liguori, Placidi, Albanese, et al., 2014; Liguori, Placidi, et al., 2016; Lozano-Tovar et al., 2024; Shimada et al., 2020; Strittmatter et al., 1996; P. Wang et al., 2021). Whereas the CSF marker deviations in narcolepsy are bidirectional, most evidence points towards lower levels in spinal CSF of amyloid-β, phosphorylated and total tau compared to healthy controls (Jennum et al., 2017; Liguori, Placidi, Albanese, et al., 2014; Liguori, Placidi, et al., 2016). It is also hypothesised that the lowest levels of amyloid-β are present at disease onset if it is triggered by inflammation, and that long-term treatment with psychostimulants might induce neuronal metabolic changes resulting in higher levels of neurodegenerative markers in the CSF (Liguori, Placidi, Izzi, et al., 2014). Whereas, lower levels of waste molecules in spinal CSF could suggest waste accumulation in the brain as a result of impaired clearance, it remains unclear whether lower marker levels in people with narcolepsy are related to altered brain clearance. Third, findings from MRI diffusivity studies in people with narcolepsy, which identified widespread diffusivity alterations, further hint towards possible fluid motion changes in narcolepsy. Specifically, a lower measure of diffusion directionality (fractional anisotropy), higher measure of axon myelination (radial diffusivity), and unaffected measures of axon organisation and mean diffusion strength (axial diffusivity, mean diffusivity) were identified in people with narcolepsy compared to controls (Gool et al., 2019; Juvodden et al., 2018; Park et al., 2016; Tezer et al., 2018). The diffusivity alterations may suggest white matter alterations of lower myelination, lower axonal density, or greater axonal diameter, but this was not fully confirmed in post-mortem histology experiments (Gool et al., *submitted*). It remains unclear to what extent myelin and axonal integrity are affected in narcolepsy type 1, or whether other factors contributed to the diffusivity differences (Gool et al., 2024). We consider the possibility that these diffusivity alterations are related to brain clearance alterations in narcolepsy, for instance through larger perivascular spaces and their contribution to the diffusivity measures.

### MRI-based surrogates of brain clearance

Neuroimaging studies have applied contrast-enhanced MRI to monitor tracer clearance and study the human brain clearance system *in vivo*. Other MRI methods, such as adaptations of arterial spin labelling and diffusion-weighted imaging, have been pursued to image brain clearance with a non-invasive approach (as summarised in van der Thiel et al., 2023).

Taoka and colleagues (Taoka et al., 2017) proposed a post-processing technique to be applied to diffusion tensor images: diffusion tensor image analysis along the perivascular space (DTI-ALPS, SFig. 1). The DTI-ALPS index aims to reflect diffusion along the perivascular space as a measure of brain clearance and has been widely applied in a diverse range of clinical populations (>140 publications in the last 5 years). The index is calculated based on diffusivity in a small area of the brain with a unique, perpendicular position of two fibre tracts and blood vessels, next to the lateral ventricles. Neurodegenerative disorders in which protein accumulations play a central role, such as Alzheimer’s and Parkinson’s disease, showed lower DTI-ALPS indices compared to healthy controls (Cai et al., 2023; Chang et al., 2023; Chen et al., 2021; Ma et al., 2021; Taoka et al., 2017). Of note, the index and what it claims to reflect has been disputed as DTI-ALPS is not CSF-nor ISF-selective and is only measured at a single location in the brain. Changes in DTI-ALPS index could therefore also originate from changes in tissue pulsatility, changes in white matter structure or presence of more fluid in the brain (i.e. enlarged PVS, free water due to demyelination, white matter hyperintensities; Ringstad, 2024; Taoka et al., 2024).

Another non-invasive method allows to investigate the coupling between neurovascular-driven fluctuations of the blood volume in the cortical grey matter with CSF oscillations at the level of the fourth ventricle (Fultz et al., 2019). An anti-correlation is observed between cortical grey matter activity (BOLD signal) and CSF waves in awake participants, which is significantly enhanced during sleep. The coupling between the two signals is explained by the following mechanism: activity fluctuations in the cortex give rise to cerebral blood volume changes, which in turn push CSF out, or allow for CSF inflow, due to the restricted space inside the skull (Monroe-Kellie doctrine). Altogether this dBOLD-CSF coupling reflects how cerebral blood volume changes can be a driving force for CSF mobility. The dBOLD-CSF coupling method has been applied in healthy people during sleep (Fultz et al., 2019), and Alzheimer’s Disease (Han, Chen, et al., 2021) and Parkinson’s Disease during wake (Han, Brown, et al., 2021; Z. Wang et al., 2023).

### MRI-based surrogates of brain clearance in narcolepsy

In narcolepsy, the DTI-ALPS index has been investigated twice. Gumeler and colleagues (2023) found no significant group differences while Hu and colleagues (2024) identified a lower DTI-ALPS index, bilaterally, in narcolepsy type 1 compared to controls. Significant correlations of the index with clinical features (polysomnography, sleepiness scales) were described in both studies. The dBOLD-CSF coupling approach has not been performed in narcolepsy, but showed stronger coupling during sleep compared to wake in healthy volunteers (Fultz et al., 2019).

Altogether, the question remains whether brain clearance is altered in narcolepsy type 1, and whether this can be reliably studied using MRI-based surrogates of brain clearance. The current study aimed to investigate changes in the DTI-ALPS index and dBOLD-CSF coupling in narcolepsy type 1. We expected to see impaired, weaker brain clearance (lower DTI-ALPS index, weaker dBOLD-CSF coupling) in people with narcolepsy type 1 compared to controls, and in relation to markers of disease severity.

## Methods

### Participants

The dataset consisted of 12 people with drug-free narcolepsy type 1 and 11 healthy, age- and sex-matched controls. Recruitment of people with narcolepsy was carried out through our tertiary sleep clinic (Sleep-Wake Centre SEIN). Participants were diagnosed with narcolepsy type 1 according to the international classification of sleep disorders, third edition (American Academy of Sleep Medicine, 2014). The polysomnography and multiple sleep latency test (MSLT) were performed for the diagnostic process and were not linked to the MRI scan. As part of regular care, CSF hypocretin-1 levels were available for nine out of 12 people with narcolepsy, and all showed to be deficient according to the narcolepsy type 1 cut-off of 110 pg/mL (Mignot et al., 2002). Three people with narcolepsy did not present with clear cataplexy and received a lumbar puncture to determine hypocretin-1 levels, which appeared deficient for two out of three. The third person showed lower levels than typical (138 pg/mL) and was still included based on the presentation of a typical clinical phenotype of narcolepsy. All participants with narcolepsy were HLA DQB1*0602-positive. Participants were excluded if they (1) used psychotropic drugs; (2) were diagnosed with other serious medical conditions; (3) showed macroscopic brain abnormalities; (4) had contraindications to receive an MRI scan. All individuals had to (1) be between 18-65 years old; (2) have normal or corrected-to-normal vision; and (3) be right-handed according to the Edinburgh Handedness Inventory (Oldfield, 1971). Medication use (n=7 drug-naïve; n=4 methylphenidate; n=1 modafinil) was interrupted for at least two weeks and no caffeine-containing substances were consumed in the 24 hours prior to the MRI session. The Dutch Adult Reading Test (proxy for intelligence; Schmand et al., 1991) and Epworth Sleepiness Scale (ESS; Johns, 1991) were administered on the day of the MRI scan. All participants signed informed consent and the study protocol was approved by the medical ethical committee of Leiden University Medical Center. Experiments were conducted in accordance with the declaration of Helsinki.

### Image acquisition

All participants were scanned using a 3T Philips Achieva MRI scanner (Philips Healthcare, Best, the Netherlands) with a 32-channel head coil and sponge cushions to minimise head motion. Diffusion images were collected with a single-shot, spin-echo EPI sequence (2×2×2⍰mm^3^ voxel size) with TR=6700⍰ms; TE=72⍰ms; FOV=224×224×120⍰mm; flip angle=90°; acquisition time=5.9 min. This protocol was executed twice with reversed k-space readout (phase-encoding in anterior-posterior, AP, and posterior-anterior, PA, direction) along 46 non-collinear directions with b=100011s/mm^2^ and b=011s/mm^2^. We acquired 3D T1-weighted images (1×1×111mm^3^) with TR=8.2⍰ms; TE=3.8⍰ms; TI=670.4⍰ms; FOV=240×240×220 mm^3^; flip angle=8°; acquisition time=6.2 min. Resting-state (rs) functional images (2.5×2.5×2.5mm3 voxel size) were acquired with TR=2250 ms; TE=29.94 ms; FOV=200×200×104.25 mm; flip angle=80°; 315 dynamics, acquisition time=11.8 min. Concurrent high-density EEG was acquired during the rs-fMRI scan to objectively verify wakefulness. EEG acquisition using carbon-wire loops and pre-processing was performed according to methods described in earlier work (van der Meer et al., 2016a, 2016b). To reduce associated artefacts, the helium pump was temporarily switched off. Cleaned EEG recordings were sleep scored according to the AASM criteria using 30-second epochs.

### Diffusion tensor image processing and DTI-ALPS index

Diffusion image corrections were performed using the FMRIB Software Library’s Diffusion Toolbox (FSL, v5.0; Andersson et al., 2003; Andersson & Sotiropoulos, 2016) as described in a previous study within the same dataset (Gool et al., 2019). Tensor fitting was run with the *--save tensor* optional argument to extract the full diffusion tensor for the purpose of this study (DTIFIT, FSL v6.0.6.4). The DTI-ALPS method was performed as described by Taoka and colleagues (2017, SFig. 1). For each participant, four 3×3 voxel ROIs were drawn on the diffusion tensor ellipsoid maps by one rater, based on the following instructions: (1) use the sagittal and transverse image to find the most superior part of (or just above) the lateral ventricle - this is where most of the deep medullary veins are located in a perpendicular position to the fibres; (2) try different levels of the sagittal plane to find a location with clear separation of the projection (blue) and association (green) fibres; (3) draw the ROIs in areas with clear head-foot (projection) and anterior-posterior (association) fibre directions based on the ellipsoid shape. No supporting images were available to better identify the deep medullary veins, such as susceptibility-weighted images. FSLutils commands were applied to extract the diffusivities in three directions of the diffusion tensor, xx, yy, and zz directions for the four ROIs (xx, yy, zz refer to the subject-specific coordinate system of the diffusion tensor). The DTI-ALPS index was calculated unilaterally for each participant, and for both images with opposing AP and PA phase-encoding directions, according to the following formula (1). We tested for differences between the left and right hemisphere, and between the AP and PA phase-encoding directions, to average in case of non-significant differences. To extend evidence in the current debate on what the DTI-ALPS index reflects, we aimed to provide quality control measures on the index, such as left-right comparisons and its relation to the dBOLD-CSF coupling amplitude. Step-by-step description of our approach and code to extract diffusivities and calculate the DTI-ALPS index are available from github.com/evavanheese/DTI-ALPS.

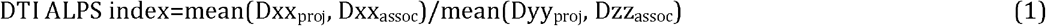

### Functional image preprocessing and dBOLD-CSF coupling

#### BOLD signal

The resting-state functional images were slice-time- and motion-corrected using the HALFpipe pipeline for harmonised fMRI analysis (Waller et al., 2022; github.com/HALFpipe/HALFpipe). HALFpipe also transformed the output images to MNI standard space, according to the most current and detailed template available (MNI152NLin2009cAsym; Horn, 2016). Cortical grey matter segmentation was performed on the T1-weighted images using FreeSurfer recon-all (FS5.3; Fischl et al., 2002). The transformation matrix from the T1-weighted image to MNI standard space was applied to the cortical grey matter mask (using ANTs registration tools) to extract the average time series from the mask in the same standard space (using FSLutils).

#### CSF signal

As there is no blood in the CSF ROIs, the signal retrieved from them is not a BOLD signal. In fact, the CSF signal in this study originates from the inflow effect, stating that fresh inflow of CSF, that has not been exposed to radiofrequency (RF) pulses flowing into a slice, has a higher signal intensity than the surrounding tissue in that slice which has been exposed to many RF pulses. With each upward slice, the fluid signal intensity decays until it reaches a steady state, similar to the surrounding tissue (Fultz et al., 2019). Therefore, edge slices are the ideal locations for the most precise measurement of CSF oscillations. Raw functional images were used to extract the signal, as motion correction can alter voxel slice position and performing such corrections correctly on edge slices is complex (Fultz et al., 2019). The first slice was rated in terms of motion and the anatomical position classified into three locations with guidance of the T1-weighted image (the CSF was either at the level of the fourth ventricle, at the level of the lower cerebellum, or within the central canal). Free-form CSF ROIs composed of connecting voxels were drawn manually on the bottom slice of the functional image and ranged from 3-11 voxels in size. The influence of the anatomical location of the CSF ROI on the dBOLD-CSF coupling amplitude was investigated in an explorative analysis. Step-by-step description of our approach and code to create a cortical grey matter mask based on FreeSurfer output, perform registrations, and extract the BOLD and CSF signal from the ROIs are available from github.com/evavanheese/BOLD-CSF.

#### Signal processing and cross-correlation

The BOLD and CSF signals were normalised, detrended, and low-pass filtered (<0.1 Hz). We then calculated the negative derivative of the BOLD signal (dBOLD), reflecting the change in BOLD signal (i.e. slope of the signal), and set negative values to zero to better reflect inflow and not outflow of CSF (Fultz et al., 2019). To cross-correlate both signals, we tested different time lags from −20 to 20 seconds and retrieved the lag and amplitude of the peak (positive or negative) closest to the lag of zero seconds. Time courses, as well as cross-correlation plots, were visually inspected for each subject to confirm a coupling of the CSF with dBOLD signal. Besides the coupling, fluctuations in CSF and dBOLD time series were further investigated by taking the area under the curve (AUC) of the CSF and dBOLD signal time courses. This experiment was done to investigate possible alterations in both signals separately, in addition to the coupling between them. Matlab scripts for signal processing and cross-correlation are available upon request.

### Covariates and statistical analysis

A preregistration of our DTI-ALPS and dBOLD-CSF analysis plan and hypotheses was submitted to the Open Science Framework prior to conducting any of the analyses (https://osf.io/mpnat). The primary outcome measures for statistical analysis in this study were: left, right, and bilateral average of the DTI-ALPS index, and dBOLD-CSF coupling amplitude and lag. Group differences and correlations with the ESS were investigated using regression models in R (R Core Team, 2014). Cohen’s *D* and Pearson’s *r* effect sizes were respectively calculated for each statistical test. All data points were investigated in relation to a range of three times the interquartile range (IQR), and classified as outliers if they exceeded this range. No outliers were identified for the primary outcome measures.

## Results

### Final sample

The data analysis was performed on 12 people with narcolepsy type 1 and 11 healthy controls. An overview of the demographic and clinical characteristics from the full sample can be found in Table 2.

### DTI-ALPS

We did not detect a significant difference in DTI-ALPS index between the left and right hemisphere (*p*=.683; SFig. 2) or AP and PA phase-encoding directions (*p*=.430). Consequently, for each participant, an average of the left and right hemisphere (*bilateral average*) was calculated for subsequent analyses. From the comparison of the bilateral average DTI-ALPS index, no significant differences were found between people with narcolepsy type 1 and matched controls (*p*=.461; Fig. 1, Table 3). The DTI-ALPS index did also not significantly correlate with ESS (*p*=.154; Fig. 4).

**Fig. 1.**
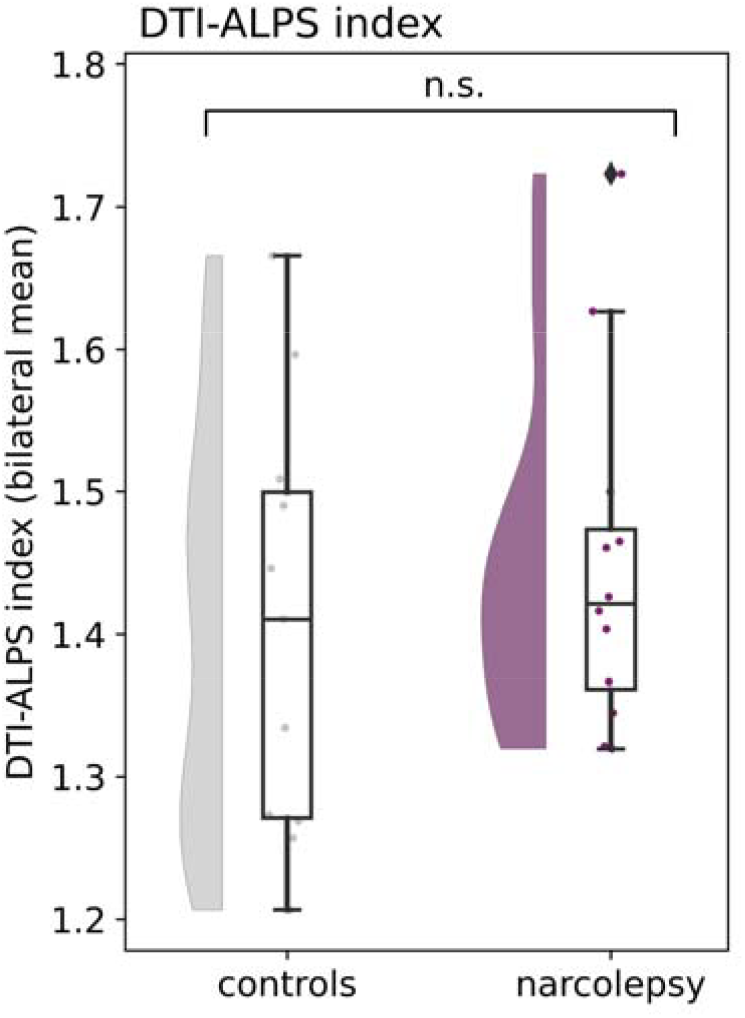
DTI-ALPS index group differences for the left and right hemisphere. No significant differences were found between the hemispheres and between narcolepsy and controls. Means, standard deviations, and estimates from the regression model are reported in Table 3.

### dBOLD-CSF coupling

We explored possible effects of CSF ROI anatomical location on the coupling strength, by rating and dividing the participant images into three categories: fourth ventricle (n=6), lower cerebellum (n=11), and central canal (n=6). The dBOLD-CSF coupling amplitudes did not differ significantly between the three locations and were pooled for the group comparison (fourth ventricle-lower cerebellum, *p*=.908; fourth ventricle-central canal, *p*=.382; lower cerebellum-central canal, *p*= .323; SFig. 4). Between people with narcolepsy type 1 and matched controls, no significant difference was found for dBOLD-CSF coupling amplitude (*p*=.263; Fig. 2, SFig. 3, Table 4). We also did not observe a significant correlation between the dBOLD-CSF coupling amplitude and the ESS (*p*=.208; Fig. 4). When investigating the dBOLD and CSF signals separately, no differences were found in the area under the curve between people with narcolepsy and controls (dBOLD, *p*=.239; and CSF, *p*=.304; Fig. 3, Table 4).

**Fig. 2.**
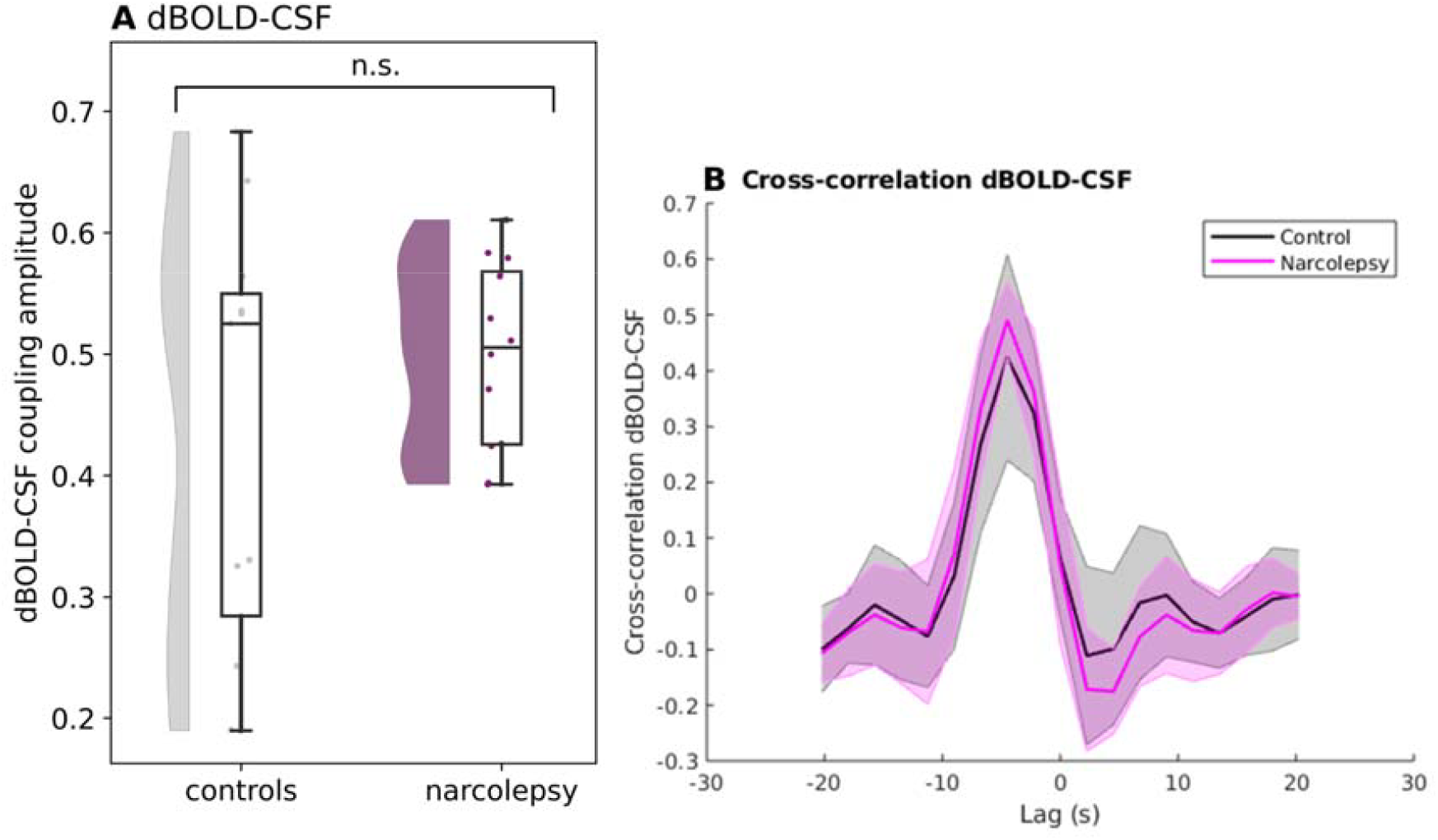
BOLD-CSF coupling amplitude (A) and coupling curve (B). (A) Quantification of retrieved amplitudes for narcolepsy (purple) and controls (grey). (B) Mean ± SD dBOLD-CSF coupling curve for narcolepsy (pink) and controls (grey). The peak closest to zero has an average amplitude and lag of 0.50 and −4.13 seconds (narcolepsy) and 0.43 and −4.09 seconds (controls). Means, standard deviations, and estimates from the regression model are reported in Table 4.

**Fig. 3.**
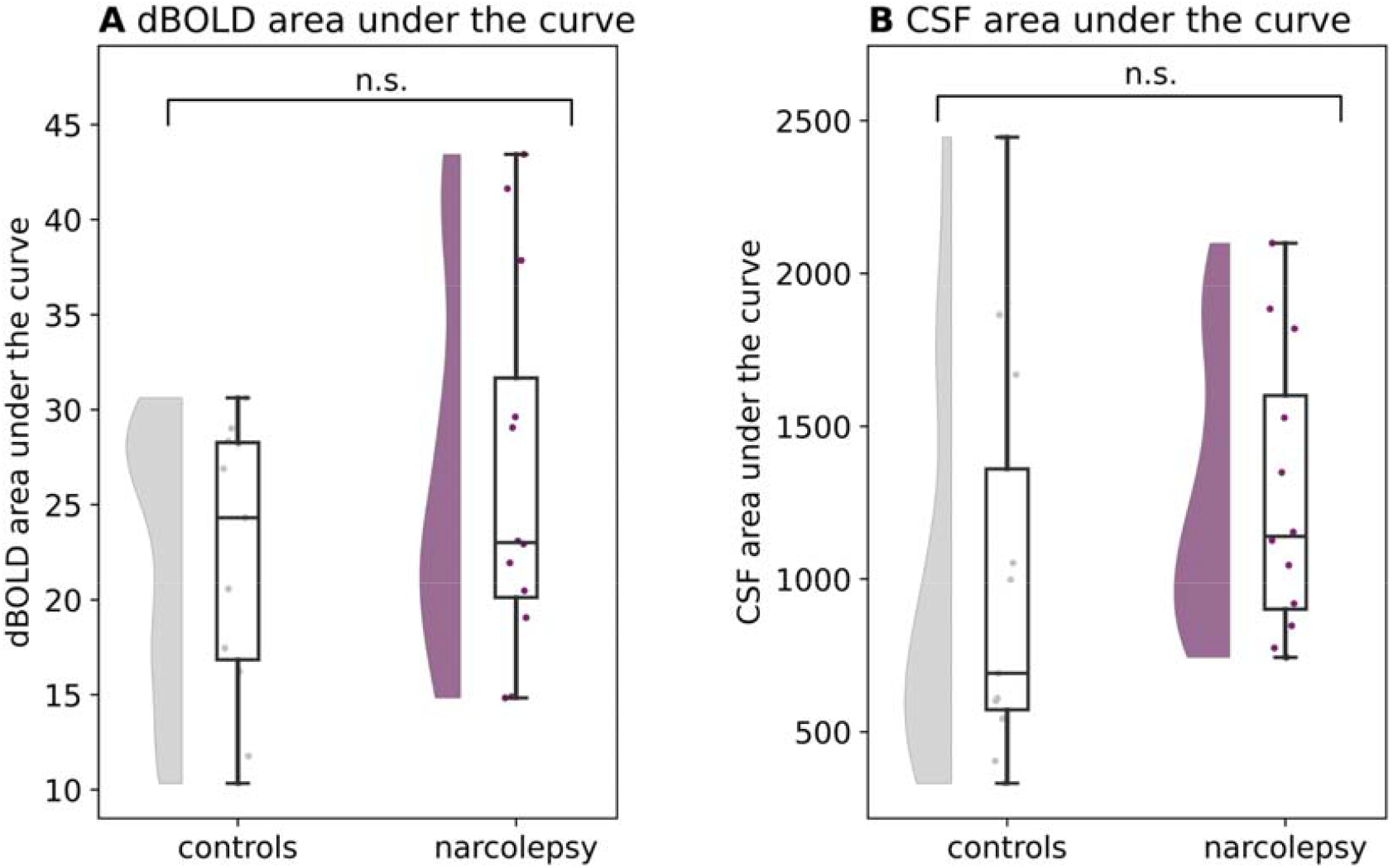
Signal fluctuations in the dBOLD (A) and CSF (B) signal. Area Under the Curve (AUC) is shown for normalised, detrended, and filtered dBOLD and CSF time series. The AUC did not significantly differ between people with narcolepsy and controls for the dBOLD (*p*=.239), as well as the CSF time series (*p*=.304).

**Fig. 4.**
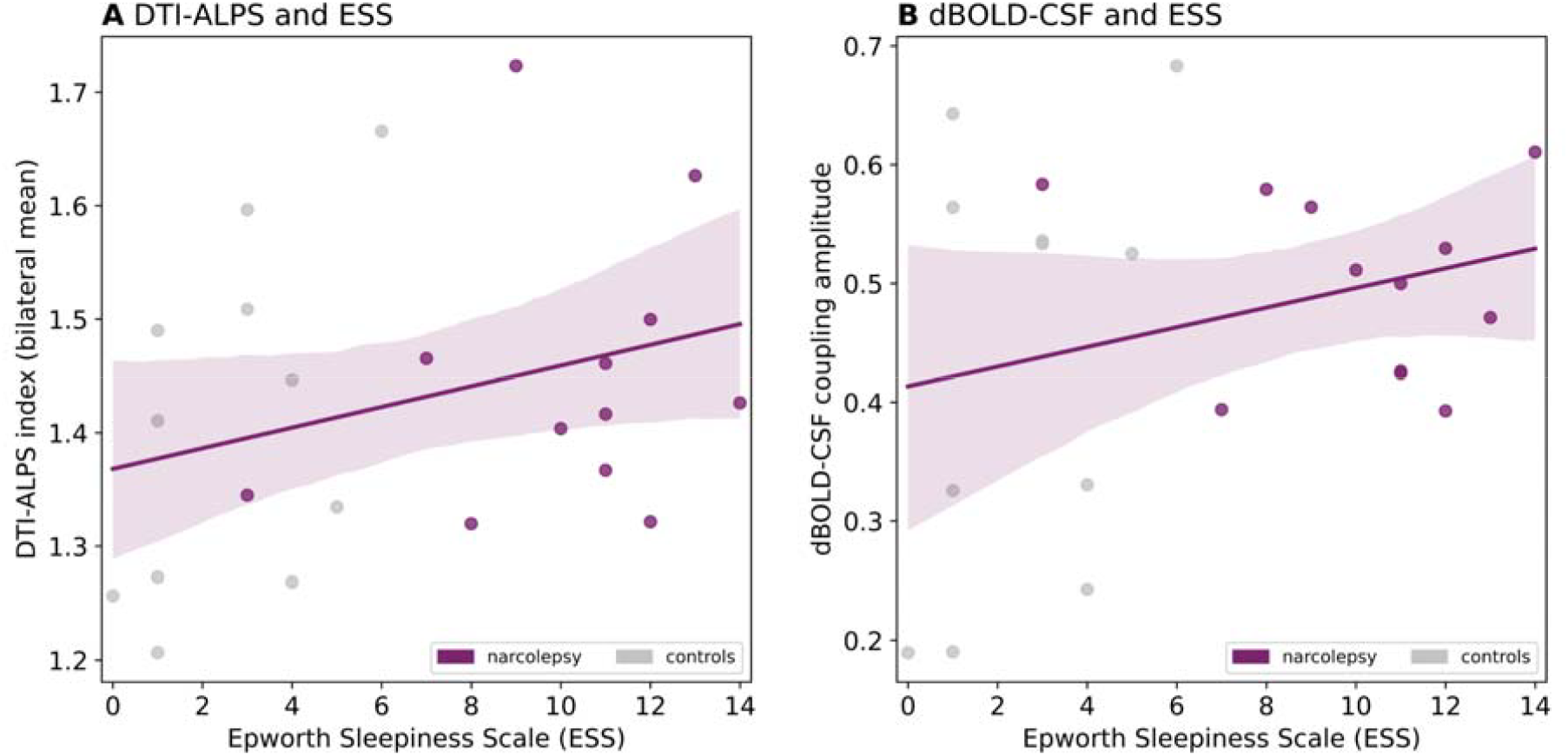
DTI-ALPS (A) and dBOLD-CSF coupling (B) correlated with Epworth Sleepiness Scale, a measure of excessive daytime sleepiness. Regression lines are drawn based on data points from narcolepsy and controls combined. DTI-ALPS: Pearson’s *r* = 0.31, *p* value = .154; BOLD-CSF: Pearson’s *r* = 0.27, *p* value = .208. Abbreviations: ESS, Epworth Sleepiness Scale.

### DTI-ALPS and dBOLD-CSF coupling

The Pearson’s correlation between the two surrogates of brain clearance appeared significant with a positive correlation coefficient (i.e. higher DTI-ALPS index relates to a higher dBOLD-CSF coupling amplitude; *p*=.025, *r*=0.46; Fig. 5).

**Fig. 5.**
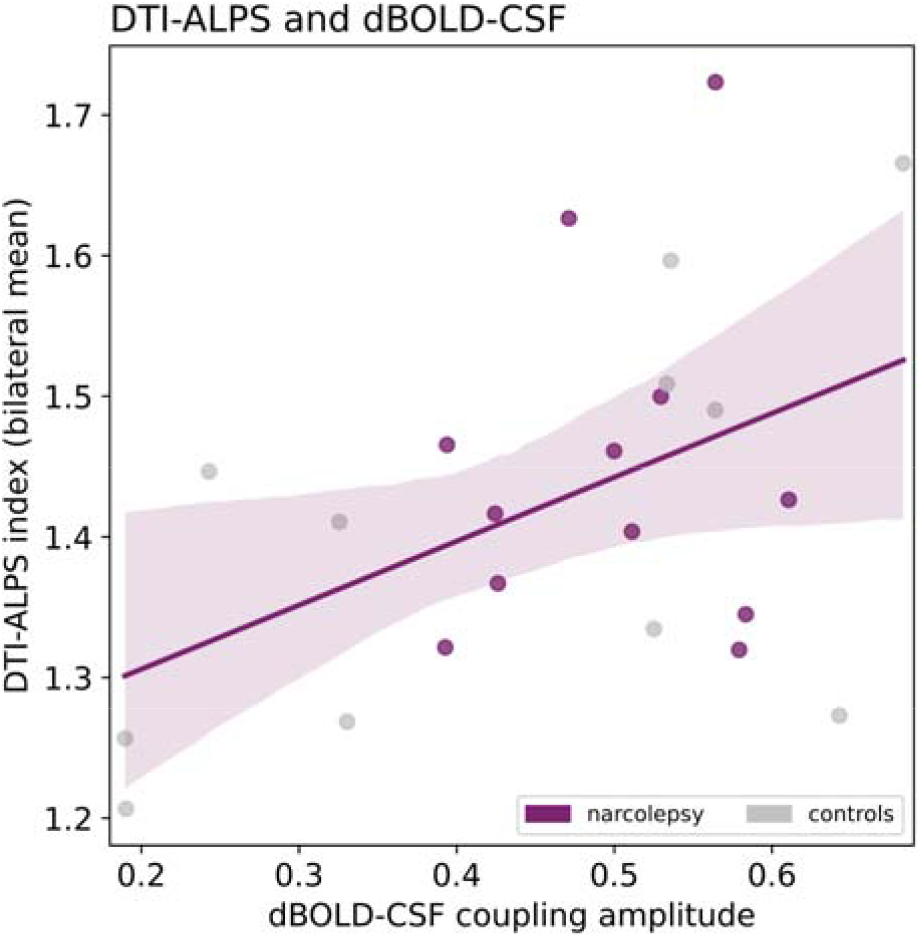
Correlation between DTI-ALPS index and dBOLD-CSF coupling amplitude. Higher DTI-ALPS index (y) is significantly correlated to larger dBOLD-CSF amplitude (x). Pearson’s *r* = 0.46, *p* value =.025.

## Discussion

In summary, we did not find significant differences between narcolepsy type 1 and healthy controls for either of the surrogate brain clearance measures, nor significant relations with excessive daytime sleepiness. The identified values were similar to previous research for both the DTI-ALPS index (here: 1.1 to 1.8; Taoka et al., 2017: 0.8 to 1.9) and dBOLD-CSF coupling amplitude (here: 0.2 to 0.7; Fultz et al., 2019: peak of averages at 0.59) and lag (here: −3.6s; Fultz et al., 2019: −1.8s), suggesting reproducibility across different study populations. The correlation between the DTI-ALPS index and dBOLD-CSF coupling amplitude was positive and significant (*p*=.025), indicating that participants with a high DTI-ALPS index were also likely to have a high dBOLD-CSF coupling amplitude, suggesting that both measures might originate from the same underlying physiology. Altogether, this study does not provide evidence for altered brain clearance in narcolepsy type 1 during wake, based on the surrogate MRI measures investigated. However, we want to emphasise that the two surrogate markers have clear limitations in their relation with brain clearance and the absence of significant differences should not be interpreted as an absence of brain clearance alterations in this population. We will discuss these limitations below in more detail.

### Altered brain clearance in narcolepsy?

#### DTI-ALPS

The absence of group differences in the DTI-ALPS index aligns with findings from one of the two previous applications of this method in narcolepsy (Gumeler et al., 2023). The authors from this study did not find between-group differences between narcolepsy type 1 and 2 combined (n=25), compared to controls (n=11). However, Hu and colleagues (2024), who performed the analysis in a larger dataset, identified a lower DTI-ALPS index bilaterally in narcolepsy type 1 (n=31) compared to controls (n=23). The DTI-ALPS index did not significantly correlate with the ESS. In the current study, the correlation between the DTI-ALPS index and ESS also appeared non-significant. Whereas the same dataset showed significant group differences on several diffusivity outcome measures in a previous investigation (Gool et al., 2019), it is likely that subtle differences in the DTI-ALPS index were not detected here due to limited statistical power.

#### dBOLD-CSF

We did not find significantly altered dBOLD-CSF coupling in narcolepsy type 1, contrary to our expectations. As our findings can only be compared to applications in other populations, it remains unclear whether dBOLD-CSF coupling is truly unaltered in narcolepsy type 1. Previous studies showed a lower coupling strength in neurodegenerative disorders (Han, Brown, et al., 2021; Han, Chen, et al., 2021; Z. Wang et al., 2023), characterised by large brain changes including protein accumulations and impaired vascular reactivity. In the absence of these significant brain alterations, that are likely linked to brain clearance, it is possible that people with narcolepsy type 1 do not show altered dBOLD-CSF coupling. Yet, as we know from previous work in healthy individuals that dBOLD-CSF coupling is stronger during sleep (Fultz et al., 2019), we speculate that people with narcolepsy might show normal coupling during wake, but altered coupling during sleep.

#### Other MRI-based measures

While the MRI-based surrogates investigated in this study have seen limited application in narcolepsy, it is worth noting that another measure has been studied that is loosely associated with brain clearance and related physiological processes. The findings from this study may provide some contextual insight, but should be interpreted with caution, as the exact role of this measure in relation to brain clearance is not yet fully understood. Järvelä and colleagues compared brain pulsations measured with ultrafast functional MRI between narcolepsy type 1 (n=23) and controls (n=23; Järvelä et al., 2022). They found a greater variance of pulsations in the low-frequency band and lower variance in the respiratory band in narcolepsy compared to controls, most prominent in the ascending arousal network. The authors propose that these results imply impaired parenchymal and ventricular CSF flow, likely related to a lack of hypocretin signalling on the downstream ascending arousal network and cortical regions.

#### Hypocretin, sleep, and amyloid-β

Both animal and human studies have indicated a relation between hypocretin signalling, sleep, and amyloid-β in the brain (Berteotti et al., 2021). Strikingly, lower levels of hypocretin, as observed in narcolepsy type 1, seem to have a neuroprotective effect. This idea is based on Alzheimer mice experiments showing that hypocretin infusion increased amyloid-β levels in the ISF, whereas hypocretin antagonists or hypocretin knockouts showed lower levels and less plaque formation (Kang et al., 2009). As these changes in amyloid-β levels co-occurred with improved or disrupted sleep, it is proposed that this effect of hypocretin on amyloid-β metabolism and clearance is solely mediated through the sleep-wake cycle (Berteotti et al., 2021). In humans, it is known that hypocretin and amyloid-β in the CSF show circadian fluctuations related to each other (Slats et al., 2012). Additionally, significantly higher levels of hypocretin are observed in CSF of people with AD pathology, and higher levels correlated with insomnia symptoms (Liguori, Nuccetelli, et al., 2016). In animals, as well as humans, beneficial effects of hypocretin antagonists have been observed on sleep, amyloid-β-related neurodegeneration, and cognition (Berteotti et al., 2021). Specifically to narcolepsy, it is proposed that the low hypocretin signalling reduces the risk of amyloid-β accumulations and thus Alzheimer’s Disease (Berteotti et al., 2021). Evidence, so far, is sparse, with one post-mortem study showing a similar prevalence of amyloid-β neuropathology as in the general population (Alzheimer’s Disease was present in 4 out of 12 cases of narcolepsy type 1; (Scammell et al., 2012) and one PET study showing a lower brain amyloid burden and lower pathological amyloid accumulation in individuals with narcolepsy (Gabelle et al., 2019).

All in all, it remains to be further explored whether brain clearance is altered in narcolepsy type 1. As the investigation of brain clearance in narcolepsy is only in its infancy, future research should aim to further explore the hypothesis of altered brain clearance in narcolepsy type 1, and consider other central disorders of hypersomnolence, and the importance of comparing different states of consciousness. With more research on brain clearance in narcolepsy type 1, we aim to determine whether brain clearance alterations are present, and if so, to better understand whether they are intrinsic to the disease, a consequence of hypocretin deficiency, or a result of disrupted sleep.

### Methodological considerations

#### Data variability

The range of data points from the two groups in this study shows a distinct pattern. For the dBOLD-CSF coupling amplitude, people with narcolepsy type 1 show a narrower range compared to the controls (Fig. 2). A similar, although less strong, pattern is observed for the CSF Area under the Curve (Fig. 3B). This pattern could originate from the narcolepsy group being very homogenous, with similar pathology and corresponding brain correlates.

#### DTI-ALPS

The DTI-ALPS method (Taoka et al., 2017) is recent, but widely applied in the past years. The main reason for this popularity is likely related to the possibility of applying the method retrospectively, on already processed DTI metrics. In addition, the methodological procedure is easy and not highly time-consuming. The validity of the DTI-ALPS index, however, has been debated, with a focus on what the index measures (Ringstad, 2024; Taoka et al., 2024). Without violation of the underlying assumptions, the index supposedly reflects perivascular fluid flow in a small region of brain parenchyma next to the lateral ventricle, in the perivascular channels surrounding the deep medullary veins. The measure is thus highly specific to a small area of the brain and does not present an accurate reflection of CSF-mediated clearance at the whole-brain level. Regional brain clearance effects, as suggested in previous studies (Eide et al., 2021), would therefore not be captured in the DTI-ALPS index. The assumed unique position of the projection and association fibres in relation to the veins, perpendicular in all directions, can also be questioned. In this study, similar to many other applications of the DTI-ALPS method, the absence of an image visualising veins (i.e. susceptibility-weighted image), led us to draw the regions of interest without confirming the presence of underlying deep medullary veins. Finally, possible violation of other assumptions, as the sequence is not CSF-nor ISF specific, could result in the DTI-ALPS index not accurately representing perivascular fluid flow, but rather residual diffusion, tissue motion, slow blood flow, or white matter alterations. The doubts surrounding the DTI-ALPS method led us to perform a multi-perspective approach in this study, which allowed for a comparison of the DTI-ALPS outcomes with the dBOLD-CSF coupling analysis.

#### dBOLD-CSF

In contrast to the DTI-ALPS method, the dBOLD-CSF analysis explores the forces driving CSF oscillations by measuring the coupling between cortical grey matter fluctuations and CSF flow at the level of the fourth ventricle. Whereas higher coupling has been observed during sleep, earlier studies confirmed that this coupling can also be observed (to a smaller extent) in awake individuals (Han, Brown, et al., 2021; Han, Chen, et al., 2021; Z. Wang et al., 2023). The dBOLD-CSF coupling represents one of the drivers of CSF flow, neurovascular activity, but in relation to CSF only measured at the level of the fourth ventricle (i.e. reflective of large scale CSF mobility which can be different from CSF-mobility close to the neurons where neuronal waste is produced). The question remains whether this measure, especially when altered in clinical populations, can be translated to provide information about aspects of clearance inside the entire brain, for example perivascular fluid flow.

#### Complementary surrogates of brain clearance

The significant positive correlation between the DTI-ALPS index and dBOLD-CSF coupling suggests that both surrogates capture (parts of) the same or similar processes. Based on the proposed mechanisms behind the measures, we hypothesise that the DTI-ALPS index and dBOLD-CSF coupling offer different perspectives of the same underlying physiology, complementing each other methodologically. Future investigations should aim to identify the exact relationships between the two measures.

#### Study set-up

The current study acquired images with a TR of 2250 ms, compared to a TR of 367 ms in the study of Fultz and colleagues (2019). A faster sampling of the functional MRI signal would offer a better temporal resolution to identify the dBOLD-CSF coupling lag and possible group differences, which was not possible in this retrospective study. One of the main limitations of the current study is its small sample size. Future work in larger datasets could point out whether the absence of brain clearance alterations in narcolepsy type 1 as reflected by the DTI-ALPS and dBOLD-CSF coupling, can be replicated. Possibly, subtle alterations would only be captured by a better-powered study. If differences in these measures are detected in future studies, an interesting focus would be to investigate and compare different subtypes of hypersomnolence disorders: narcolepsy type 1, type 2, and idiopathic hypersomnia. Additionally, to gain insights into the influence of sleep on the available surrogates, future investigations could acquire MR images at different times of day, during different states of consciousness (wake, REM and NREM stages) and after sleep deprivation. Finally, other MRI-based surrogates of brain clearance could be explored in narcolepsy. Studies have attempted to estimate CSF and ISF flow or exchange using: phase-contrast MRI (Kim et al., 2016; Korbecki et al., 2019); variants of arterial spin labelling (ASL) (Petitclerc et al., 2021; Taoka & Naganawa, 2020); diffusion-based imaging (Örzsik et al., 2023; Shao et al., 2019; Wong et al., 2020; Zhang et al., 2012), motion-prepared imaging (Harrison et al., 2018; Hirschler et al., n.d., 2019) or magnetisation transfer ASL (Li et al., 2022). These measures potentially capture a different aspect, which could improve our understanding of whether and how brain clearance is affected or involved in narcolepsy type 1.

## Conclusion

To conclude, the hypothesis of altered brain clearance in narcolepsy type 1 is not supported by evidence from the current study. Future work should aim to investigate other (surrogate) measures of brain clearance to better understand the possible role of altered brain clearance in narcolepsy type 1, especially during non-wake conscious states and in comparison to other central disorders of hypersomnolence.

## Acknowledgements

We thank Alexander Leemans and Dennis Kies for their guidance during the DTI analysis, as published in (Gool et al., 2019). The DTI study formed the foundation for the DTI-ALPS analysis described here. We also thank Emiel Roefs for his advice on the image registrations.

## Author contributions

J.K.G, G.J.L, Y.D.W, and R.F, were involved in data collection. J.K.G pre-processed and analysed the diffusion MRI data for a previous study. E.M.H. further analysed the diffusion MRI data and pre-processed and analysed the functional MRI data for this manuscript, under supervision of L.H. Both J.K.G. and M.J.P.O. provided feedback during the analyses. E.M.H wrote the original draft, J.K.G, G.J.L., Y.D.W., M.J.P.O., R.F., and L.H. edited the manuscript.

## Data availability statement

The raw data underlying this article cannot be shared publicly to guarantee the privacy of individuals that participated in the study. Derived data (i.e. outcomes after diffusion/functional MRI processing) will be shared on reasonable request to the corresponding author. The processing pipelines are openly available through github.com/evavanheese/.

## Ethics approval and patient consent statement

All participants signed informed consent and the study protocol was approved by the medical ethical committee of Leiden University Medical Center. Experiments were conducted in accordance with the declaration of Helsinki.

## Permission to reproduce material from other sources

Some figures in this manuscript contain images adapted from existing publications. Specifically:

- In Supplementary Figure 1, the top left image is adapted from Kuijf et al. (2016) ‘Quantification of deep medullary veins at 7 T brain MRI,’ published in European Radiology under the terms of the Creative Commons CC-BY-NC licence.
- In Supplementary Figure 4, panel B includes an anatomical image adapted from OpenStax Anatomy and Physiology, licensed under CC-BY 4.0, with modifications.

These adaptations have been made in accordance with the terms of the respective licences.

## References

American Academy of Sleep Medicine. (2014). International classification of sleep disorders—third edition (ICSD-3). AASM Resour Libr, 281, 2313.

Andersson, J. L. R., Skare, S., & Ashburner, J. (2003). How to correct susceptibility distortions in spin-echo echo-planar images: application to diffusion tensor imaging. NeuroImage, 20(2), 870–888.

Andersson, J. L. R., & Sotiropoulos, S. N. (2016). An integrated approach to correction for off-resonance effects and subject movement in diffusion MR imaging. NeuroImage, 125, 1063–1078.

Baiardi, S., Pizza, F., Polischi, B., Moresco, M., Abu-Rumeileh, S., Plazzi, G., & Parchi, P. (2020). Cerebrospinal fluid biomarkers of neurodegeneration in narcolepsy type 1. Sleep, 43(2). 10.1093/sleep/zsz215

Bakker, E. N. T. P., Naessens, D. M. P., & VanBavel, E. (2019). Paravascular spaces: entry to or exit from the brain? Experimental Physiology, 104(7), 1013–1017.

Berteotti, C., Liguori, C., & Pace, M. (2021). Dysregulation of the orexin/hypocretin system is not limited to narcolepsy but has far-reaching implications for neurological disorders. The European Journal of Neuroscience, 53(4), 1136–1154.

Cai, X., Chen, Z., He, C., Zhang, P., Nie, K., Qiu, Y., Wang, L., Wang, L., Jing, P., & Zhang, Y. (2023). Diffusion along perivascular spaces provides evidence interlinking compromised glymphatic function with aging in Parkinson’s disease. CNS Neuroscience & Therapeutics, 29(1), 111–121.

Carare, R. O., Bernardes-Silva, M., Newman, T. A., Page, A. M., Nicoll, J. A. R., Perry, V. H., & Weller, R. O. (2008). Solutes, but not cells, drain from the brain parenchyma along basement membranes of capillaries and arteries: significance for cerebral amyloid angiopathy and neuroimmunology. Neuropathology and Applied Neurobiology, 34(2), 131–144.

Chang, H.-I., Huang, C.-W., Hsu, S.-W., Huang, S.-H., Lin, K.-J., Ho, T.-Y., Ma, M.-C., Hsiao, W.-C., & Chang, C.-C. (2023). Gray matter reserve determines glymphatic system function in young-onset Alzheimer’s disease: Evidenced by DTI-ALPS and compared with age-matched controls. Psychiatry and Clinical Neurosciences, 77(7), 401–409.

Chen, H.-L., Chen, P.-C., Lu, C.-H., Tsai, N.-W., Yu, C.-C., Chou, K.-H., Lai, Y.-R., Taoka, T., & Lin, W.-C. (2021). Associations among Cognitive Functions, Plasma DNA, and Diffusion Tensor Image along the Perivascular Space (DTI-ALPS) in Patients with Parkinson’s Disease. Oxidative Medicine and Cellular Longevity, 2021, 4034509.

Demiral, Ş.B., Tomasi, D., Sarlls, J., Lee, H., Wiers, C. E., Zehra, A., Srivastava, T., Ke, K., Shokri-Kojori, E., Freeman, C. R., & Others. (2019). Apparent diffusion coefficient changes in human brain during sleep--Does it inform on the existence of a glymphatic system? NeuroImage, 185, 263–273.

Ebrahim, I. O., Howard, R. S., Kopelman, M. D., Sharief, M. K., & Williams, A. J. (2002). The Hypocretin/Orexin System. Journal of the Royal Society of Medicine, 95(5), 227–230.

Eide, P. K., Vinje, V., Pripp, A. H., Mardal, K.-A., & Ringstad, G. (2021). Sleep deprivation impairs molecular clearance from the human brain. Brain: A Journal of Neurology, 144(3), 863–874.

Fischl, B., Salat, D. H., Busa, E., Albert, M., Dieterich, M., Haselgrove, C., van der Kouwe, A., Killiany, R., Kennedy, D., Klaveness, S., Montillo, A., Makris, N., Rosen, B., & Dale, A. M. (2002). Whole brain segmentation: automated labeling of neuroanatomical structures in the human brain. Neuron, 33(3), 341–355.

Fultz, N. E., Bonmassar, G., Setsompop, K., Stickgold, R. A., Rosen, B. R., Polimeni, J. R., & Lewis, L. D. (2019). Coupled electrophysiological, hemodynamic, and cerebrospinal fluid oscillations in human sleep. Science, 366(6465), 628–631.

Gabelle, A., Jaussent, I., Bouallègue, F. B., Lehmann, S., Lopez, R., Barateau, L., Grasselli, C., Pesenti, C., de Verbizier, D., Béziat, S., Mariano-Goulart, D., Carlander, B., Dauvilliers, Y., Alzheimer’s Disease Neuroimaging Initiative, & Multi-Domain Intervention Alzheimer’s Prevention Trial study groups. (2019). Reduced brain amyloid burden in elderly patients with narcolepsy type 1. Annals of Neurology, 85(1), 74–83.

Gool, J. K., Dang-Vu, T. T., & van der Werf, Y. D. (2024). White matter integrity in narcolepsy: the structural blueprint for functional complaints? Sleep, 47(6). 10.1093/sleep/zsae020

Gool, J. K., De Brouwer, A. C. E., Shan, L., Bol, J. G. J. M., Hoogendoorn, A. W., Van der Werf, Y. D., Lammers, G. J., Jonkman, L. E., Fronczek, R., & Schenk, G. J (2024). Widespread white matter axonal loss in narcolepsy type 1. Submitted for publication.

Gool, J. K., Fronczek, R., Leemans, A., Kies, D. A., Lammers, G. J., & Van der Werf, Y. D. (2019). Widespread white matter connectivity abnormalities in narcolepsy type 1: A diffusion tensor imaging study. NeuroImage. Clinical, 24, 101963.

Gumeler, E., Aygun, E., Tezer, F. I., Saritas, E. U., & Oguz, K. K. (2023). Assessment of glymphatic function in narcolepsy using DTI-ALPS index. Sleep Medicine, 101, 522–527.

Han, F., Brown, G. L., Zhu, Y., Belkin-Rosen, A. E., Lewis, M. M., Du, G., Gu, Y., Eslinger, P. J., Mailman, R. B., Huang, X., & Liu, X. (2021). Decoupling of Global Brain Activity and Cerebrospinal Fluid Flow in Parkinson’s Disease Cognitive Decline. Movement Disorders: Official Journal of the Movement Disorder Society, 36(9), 2066–2076.

Han, F., Chen, J., Belkin-Rosen, A., Gu, Y., Luo, L., Buxton, O. M., Liu, X., & the Alzheimer’s Disease Neuroimaging Initiative. (2021). Reduced coupling between cerebrospinal fluid flow and global brain activity is linked to Alzheimer disease–related pathology. PLoS Biology, 19(6), e3001233.

Harrison, I. F., Siow, B., Akilo, A. B., Evans, P. G., Ismail, O., Ohene, Y., Nahavandi, P., Thomas, D. L., Lythgoe, M. F., & Wells, J. A. (2018). Non-invasive imaging of CSF-mediated brain clearance pathways via assessment of perivascular fluid movement with diffusion tensor MRI. eLife, 7. 10.7554/eLife.34028

Heier, M. S., Skinningsrud, A., Paus, E., & Gautvik, K. M. (2014). Increased cerebrospinal fluid levels of nerve cell biomarkers in narcolepsy with cataplexy. Sleep Medicine, 15(6), 614–618.

Hirschler, L., Aldea, R., Petitclerc, L., Ronen, I. A., de Koning, P. J. H., van Buchem, M. A., & van Osch, M. J. P. (2019). High resolution T2-prepared MRI enables non-invasive assessment of CSF flow in perivascular spaces of the human brain. Proceedings of the 27th Annual Meeting of ISMRM, Montreal, Quebec, Canada, 746. https://archive.ismrm.org/2019/0746.html

Hirschler, L., Runderkamp, B. A., Franklin, S. L., & van Harten, T. (n.d.). The driving force of glymphatics: influence of the cardiac cycle in perivascular spaces in humans. Proceedings of the 28th.

Horn, A. (2016). Mni t1 6thgen nlin to mni 2009b nlin ants transform. Jul.

Hu, P., Zou, Y., Dai, J., Xiong, R., Zhu, H., Li, C., & Tang, X. P. (2024). Diffusion tensor imaging analysis along the perivascular space revealed the dysfunction of glymphatic system in narcolepsy type 1 patients. ISMRxM 2024.

Iliff, J. J., Wang, M., Liao, Y., Plogg, B. A., Peng, W., Gundersen, G. A., Benveniste, H., Vates, G. E., Deane, R., Goldman, S. A., Nagelhus, E. A., & Nedergaard, M. (2012). A paravascular pathway facilitates CSF flow through the brain parenchyma and the clearance of interstitial solutes, including amyloid β. Science Translational Medicine, 4(147), 147ra111.

Järvelä, M., Kananen, J., Korhonen, V., Huotari, N., Ansakorpi, H., & Kiviniemi, V. (2022). Increased very low frequency pulsations and decreased cardiorespiratory pulsations suggest altered brain clearance in narcolepsy. Communication & Medicine, 2, 122.

Jennum, P. J., Ibsen, R., Petersen, E. R., Knudsen, S., & Kjellberg, J. (2012). Health, social, and economic consequences of narcolepsy: a controlled national study evaluating the societal effect on patients and their partners. Sleep Medicine, 13(8), 1086–1093.

Jennum, P. J., Østergaard Pedersen, L., Czarna Bahl, J. M., Modvig, S., Fog, K., Holm, A., Rahbek Kornum, B., & Gammeltoft, S. (2017). Cerebrospinal Fluid Biomarkers of Neurodegeneration Are Decreased or Normal in Narcolepsy. Sleep, 40(1). 10.1093/sleep/zsw006

Johns, M. W. (1991). A new method for measuring daytime sleepiness: the Epworth sleepiness scale. Sleep, 14(6), 540–545.

Juvodden, H. T., Alnæs, D., Lund, M. J., Agartz, I., Andreassen, O. A., Dietrichs, E., Thorsby, P. M., Westlye, L. T., & Knudsen, S. (2018). Widespread white matter changes in post-H1N1 patients with narcolepsy type 1 and first-degree relatives. Sleep, 41(10). 10.1093/sleep/zsy145

Kallweit, U., Hidalgo, H., Engel, A., Baumann, C. R., Bassetti, C. L., & Dahmen, N. (2012). Post H1N1 vaccination narcolepsy-cataplexy with decreased CSF beta-amyloid. Sleep Medicine, 13(3), 323.

Kang, J.-E., Lim, M. M., Bateman, R. J., Lee, J. J., Smyth, L. P., Cirrito, J. R., Fujiki, N., Nishino, S., & Holtzman, D. M. (2009). Amyloid-beta dynamics are regulated by orexin and the sleep-wake cycle. Science, 326(5955), 1005–1007.

Kim, K. H., Choi, S. H., & Park, S.-H. (2016). Feasibility of Quantifying Arterial Cerebral Blood Volume Using Multiphase Alternate Ascending/Descending Directional Navigation (ALADDIN). PloS One, 11(6), e0156687.

Korbecki, A., Zimny, A., Podgórski, P., Sąsiadek, M., & Bladowska, J. (2019). Imaging of cerebrospinal fluid flow: fundamentals, techniques, and clinical applications of phase-contrast magnetic resonance imaging. Polish Journal of Radiology / Polish Medical Society of Radiology, 84, e240–e250.

Li, A. M., Chen, L., Liu, H., Li, Y., Duan, W., & Xu, J. (2022). Age-dependent cerebrospinal fluid-tissue water exchange detected by magnetization transfer indirect spin labeling MRI. Magnetic Resonance in Medicine: Official Journal of the Society of Magnetic Resonance in Medicine / Society of Magnetic Resonance in Medicine, 87(5), 2287–2298.

Liguori, C., Nuccetelli, M., Izzi, F., Sancesario, G., Romigi, A., Martorana, A., Amoroso, C., Bernardini, S., Marciani, M. G., Mercuri, N. B., & Placidi, F. (2016). Rapid eye movement sleep disruption and sleep fragmentation are associated with increased orexin-A cerebrospinal-fluid levels in mild cognitive impairment due to Alzheimer’s disease. Neurobiology of Aging, 40, 120–126.

Liguori, C., Placidi, F., Albanese, M., Nuccetelli, M., Izzi, F., Marciani, M. G., Mercuri, N. B., Bernardini, S., & Romigi, A. (2014). CSF beta-amyloid levels are altered in narcolepsy: a link with the inflammatory hypothesis? Journal of Sleep Research, 23(4), 420–424.

Liguori, C., Placidi, F., Izzi, F., Albanese, M., Nuccetelli, M., Bernardini, S., Marciani, M. G., Mercuri, N. B., & Romigi, A. (2014). May CSF beta-amyloid and tau proteins levels be influenced by long treatment duration and stable medication in narcolepsy? Sleep Medicine, 15(11), 1424.

Liguori, C., Placidi, F., Izzi, F., Nuccetelli, M., Bernardini, S., Sarpa, M. G., Cum, F., Marciani, M. G., Mercuri, N. B., & Romigi, A. (2016). Beta-amyloid and phosphorylated tau metabolism changes in narcolepsy over time. Sleep & Breathing = Schlaf & Atmung, 20(1), 277–283; discussion 283.

Lozano-Tovar, S., Nuccetelli, M., Placidi, F., Izzi, F., Sancesario, G., Bernardini, S., Biagio Mercuri, N., & Liguori, C. (2024). CSF dynamics of orexin and β-amyloid42 levels in narcolepsy and Alzheimer’s disease patients: A controlled study. Neuroscience Letters, 137914.

Ma, X., Li, S., Li, C., Wang, R., Chen, M., Chen, H., & Su, W. (2021). Diffusion Tensor Imaging Along the Perivascular Space Index in Different Stages of Parkinson’s Disease. Frontiers in Aging Neuroscience, 13, 773951.

Mignot, E., Lammers, G. J., Ripley, B., Okun, M., Nevsimalova, S., Overeem, S., Vankova, J., Black, J., Harsh, J., Bassetti, C., Schrader, H., & Nishino, S. (2002). The role of cerebrospinal fluid hypocretin measurement in the diagnosis of narcolepsy and other hypersomnias. Archives of Neurology, 59(10), 1553–1562.

Nishino, S., Ripley, B., Overeem, S., Lammers, G. J., & Mignot, E. (2000). Hypocretin (orexin) deficiency in human narcolepsy. The Lancet, 355(9197), 39–40.

Oldfield, R. C. (1971). The assessment and analysis of handedness: the Edinburgh inventory. Neuropsychologia, 9(1), 97–113.

Örzsik, B., Palombo, M., Asllani, I., Dijk, D.-J., Harrison, N. A., & Cercignani, M. (2023). Higher order diffusion imaging as a putative index of human sleep-related microstructural changes and glymphatic clearance. NeuroImage, 274, 120124.

Park, Y. K., Kwon, O.-H., Joo, E. Y., Kim, J.-H., Lee, J. M., Kim, S. T., & Hong, S. B. (2016). White matter alterations in narcolepsy patients with cataplexy: tract-based spatial statistics. Journal of Sleep Research, 25(2), 181–189.

Petitclerc, L., Hirschler, L., Wells, J. A., Thomas, D. L., van Walderveen, M. A. A., van Buchem, M. A., & van Osch, M. J. P. (2021). Ultra-long-TE arterial spin labeling reveals rapid and brain-wide blood-to-CSF water transport in humans. NeuroImage, 245, 118755.

Pizza, F., Vandi, S., Iloti, M., Franceschini, C., Liguori, R., Mignot, E., & Plazzi, G. (2015). Nocturnal Sleep Dynamics Identify Narcolepsy Type 1. Sleep, 38(8), 1277–1284.

R Core Team. (2014). RA Language and Environment for Statistical Computing (R Foundation for Statistical Computing).

Ringstad, G. (2024). Glymphatic imaging: a critical look at the DTI-ALPS index. Neuroradiology, 66(2), 157–160.

Roth, T., Dauvilliers, Y., Mignot, E., Montplaisir, J., Paul, J., Swick, T., & Zee, P. (2013). Disrupted nighttime sleep in narcolepsy. Journal of Clinical Sleep Medicine: JCSM: Official Publication of the American Academy of Sleep Medicine, 9(9), 955–965.

Scammell, T. E., Matheson, J. K., Honda, M., Thannickal, T. C., & Siegel, J. M. (2012). Coexistence of narcolepsy and Alzheimer’s disease. Neurobiology of Aging, 33(7), 1318–1319.

Schmand, B., Bakker, D., Saan, R., & Louman, J. (1991). [The Dutch Reading Test for Adults: a measure of premorbid intelligence level]. Tijdschrift voor gerontologie en geriatrie, 22(1), 15–19.

Shao, X., Ma, S. J., Casey, M., D’Orazio, L., Ringman, J. M., & Wang, D. J. J. (2019). Mapping water exchange across the blood-brain barrier using 3D diffusion-prepared arterial spin labeled perfusion MRI. Magnetic Resonance in Medicine: Official Journal of the Society of Magnetic Resonance in Medicine / Society of Magnetic Resonance in Medicine, 81(5), 3065–3079.

Shimada, M., Miyagawa, T., Kodama, T., Toyoda, H., Tokunaga, K., & Honda, M. (2020). Metabolome analysis using cerebrospinal fluid from narcolepsy type 1 patients. Sleep, 43(11). 10.1093/sleep/zsaa095

Slats, D., Claassen, J. A. H. R., Lammers, G. J., Melis, R. J., Verbeek, M. M., & Overeem, S. (2012). Association between hypocretin-1 and amyloid-β42 cerebrospinal fluid levels in Alzheimer’s disease and healthy controls. Current Alzheimer Research, 9(10), 1119– 1125.

Strittmatter, M., Isenberg, E., Grauer, M. T., Hamann, G., & Schimrigk, K. (1996). CSF substance P somatostatin and monoaminergic transmitter metabolites in patients with narcolepsy. Neuroscience Letters, 218(2), 99–102.

Taoka, T., Ito, R., Nakamichi, R., Nakane, T., Kawai, H., & Naganawa, S. (2024). Diffusion Tensor Image Analysis ALong the Perivascular Space (DTI-ALPS): Revisiting the Meaning and Significance of the Method. Magnetic Resonance in Medical Sciences: MRMS: An Official Journal of Japan Society of Magnetic Resonance in Medicine, 23(3), 268–290.

Taoka, T., Masutani, Y., Kawai, H., Nakane, T., Matsuoka, K., Yasuno, F., Kishimoto, T., & Naganawa, S. (2017). Evaluation of glymphatic system activity with the diffusion MR technique: diffusion tensor image analysis along the perivascular space (DTI-ALPS) in Alzheimer’s disease cases. Japanese Journal of Radiology, 35(4), 172–178.

Taoka, T., & Naganawa, S. (2020). Glymphatic imaging using MRI. Journal of Magnetic Resonance Imaging: JMRI, 51(1), 11–24.

Tezer, F. I., Erdal, A., Gumusyayla, S., Has, A. C., Gocmen, R., & Oguz, K. K. (2018). Differences in diffusion tensor imaging changes between narcolepsy with and without cataplexy. Sleep Medicine, 52, 128–133.

Thannickal, T. C., Moore, R. Y., Nienhuis, R., Ramanathan, L., Gulyani, S., Aldrich, M., Cornford, M., & Siegel, J. M. (2000). Reduced number of hypocretin neurons in human narcolepsy. Neuron, 27(3), 469–474.

van der Meer, J. N., Pampel, A., van Someren, E. J. W., Ramautar, J. R., van der Werf, Y. D., Gomez-Herrero, G., Lepsien, J., Hellrung, L., Hinrichs, H., Möller, H. E., & Walter, M. (2016a). Carbon-wire loop based artifact correction outperforms post-processing EEG/fMRI corrections—A validation of a real-time simultaneous EEG/fMRI correction method. NeuroImage, 125, 880–894.

van der Meer, J. N., Pampel, A., van Someren, E. J. W., Ramautar, J., van der Werf, Y., Gomez-Herrero, G., Lepsien, J., Hellrung, L., Hinrichs, H., Möller, H., & Walter, M. (2016b). “Eyes Open - Eyes Closed” EEG/fMRI data set including dedicated “Carbon Wire Loop” motion detection channels. Data in Brief, 7, 990–994.

van der Thiel, M. M., Backes, W. H., Ramakers, I. H. G. B., & Jansen, J. F. A. (2023). Novel developments in non-contrast enhanced MRI of the perivascular clearance system: What are the possibilities for Alzheimer’s disease research? Neuroscience and Biobehavioral Reviews, 144, 104999.

Waller, L., Erk, S., Pozzi, E., & Toenders, Y. J. (2022). ENIGMA HALFpipe: Interactive, reproducible, and efficient analysis for resting-state and task-based fMRI data. Human Brain Mapping. https://onlinelibrary.wiley.com/doi/abs/10.1002/hbm.25829

Wang, P., Li, Q., Dong, X., An, H., Li, J., Zhao, L., Yan, H., Aritake, K., Huang, Z., Strohl, K. P., Urade, Y., Zhang, J., & Han, F. (2021). Lipocalin-type prostaglandin D synthase levels increase in patients with narcolepsy and idiopathic hypersomnia. Sleep, 44(4). 10.1093/sleep/zsaa234

Wang, Z., Song, Z., Zhou, C., Fang, Y., Gu, L., Yang, W., Gao, T., Si, X., Liu, Y., Chen, Y., Guan, X., Guo, T., Wu, J., Bai, X., Zhang, M., Zhang, B., & Pu, J. (2023). Reduced coupling of global brain function and cerebrospinal fluid dynamics in Parkinson’s disease. Journal of Cerebral Blood Flow and Metabolism: Official Journal of the International Society of Cerebral Blood Flow and Metabolism, 43(8), 1328–1339.

Wong, S. M., Backes, W. H., Drenthen, G. S., Zhang, C. E., Voorter, P. H. M., Staals, J., van Oostenbrugge, R. J., & Jansen, J. F. A. (2020). Spectral Diffusion Analysis of Intravoxel Incoherent Motion MRI in Cerebral Small Vessel Disease. Journal of Magnetic Resonance Imaging: JMRI, 51(4), 1170–1180.

Xie, L., Kang, H., Xu, Q., Chen, M. J., Liao, Y., Thiyagarajan, M., O’Donnell, J., Christensen, D. J., Nicholson, C., Iliff, J. J., Takano, T., Deane, R., & Nedergaard, M. (2013). Sleep drives metabolite clearance from the adult brain. Science, 342(6156), 373–377.

Zhang, H., Schneider, T., Wheeler-Kingshott, C. A., & Alexander, D. C. (2012). NODDI: practical in vivo neurite orientation dispersion and density imaging of the human brain. NeuroImage, 61(4), 1000–1016.

Zhao, L., Tannenbaum, A., Bakker, E. N. T. P., & Benveniste, H. (2022). Physiology of Glymphatic Solute Transport and Waste Clearance from the Brain. Physiology, 37(6), 0.

