## Supplemental Figures for "MRI-based surrogates of brain clearance in narcolepsy type 1"

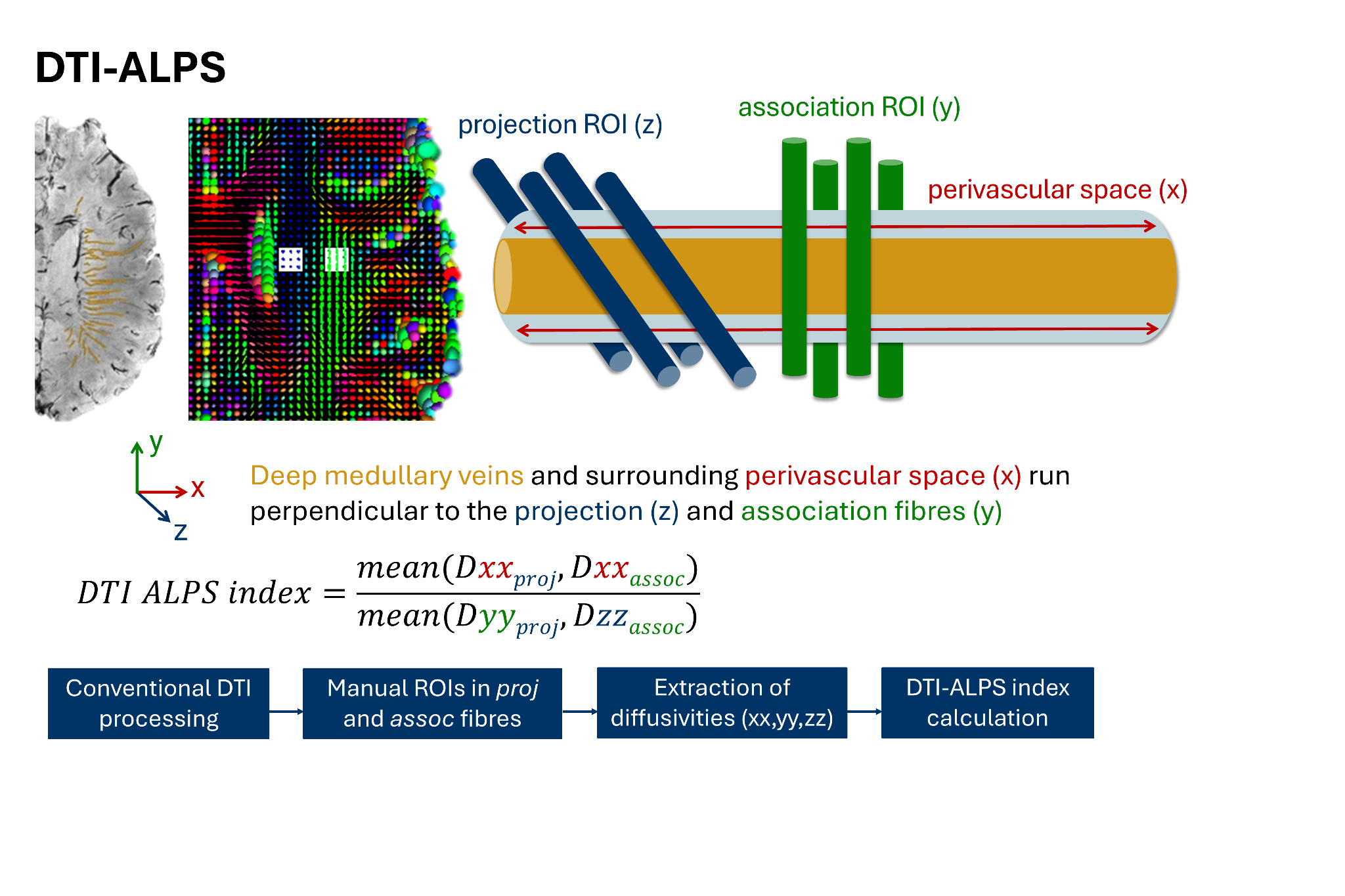

**SFig. 1. Schematic visualisation methodological concept and steps DTI-ALPS index.** From left to right, the image shows the deep medullary veins (orange) next to the lateral ventricles; the regions of interest drawn in the projection fibres (blue) and association fibres (green), on the ellipsoid map showing the DTI tensor; a schematic enlargement of the vein with projection and association fibres perpendicular to the vessel. The general analysis flow is presented in blue boxes at the bottom of the image. Note: the susceptibility-weighted image is for visualisation purposes only and was not used in the analysis. The top left image is adapted from Kuijf et al. (2016) “Quantification of deep medullary veins at 7 T brain MRI”, *Eur. Radiol.*, under the terms of the Creative Commons CC-BY-NC licence.

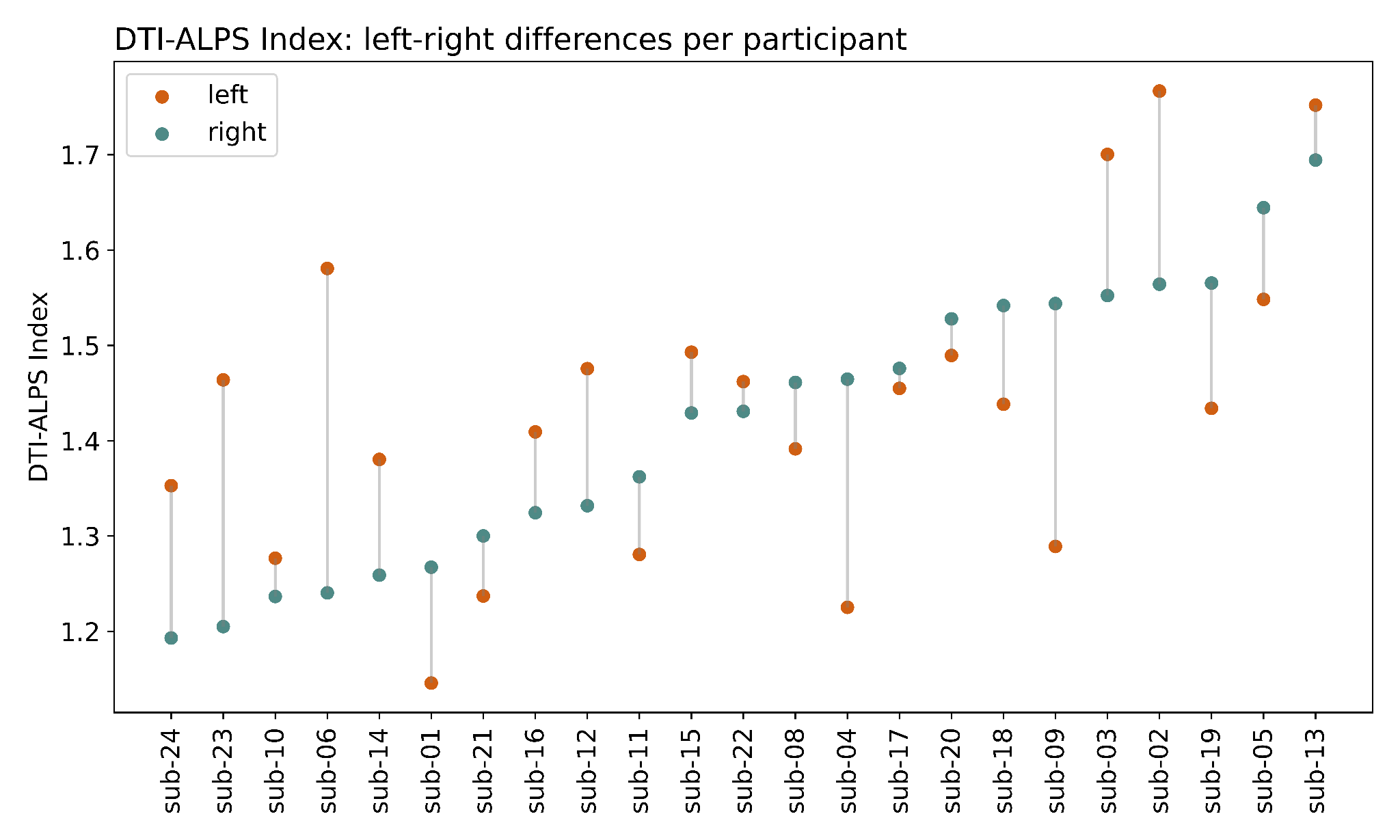

**SFig. 2. Differences in DTI-ALPS index between the left and right hemisphere.** The index is an average of the AP and PA phase-encoding direction.

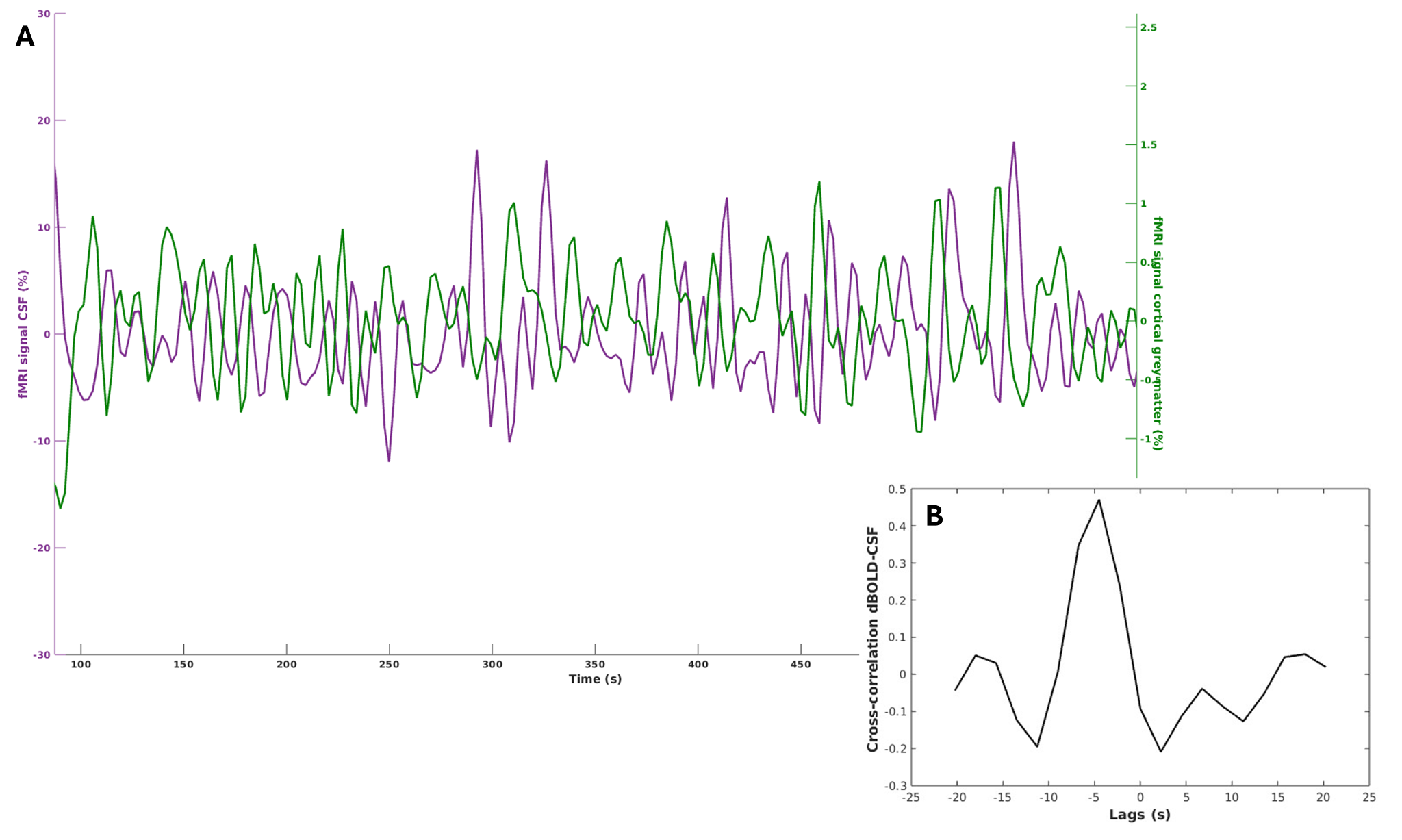

**SFig. 3. BOLD-CSF time course (A) and cross-correlation (B).** **(A)** fMRI time courses for CSF (purple) and cortical BOLD (green) from one participant (sub-03). Signals are normalised, detrended and filtered. Note: the time courses are anticorrelated at most instances where the BOLD signal decreases and the CSF increases. **(B)** Cross-correlation curve from the same participant. The peak closest to 0 has an amplitude 0.47 at -4.5 seconds.

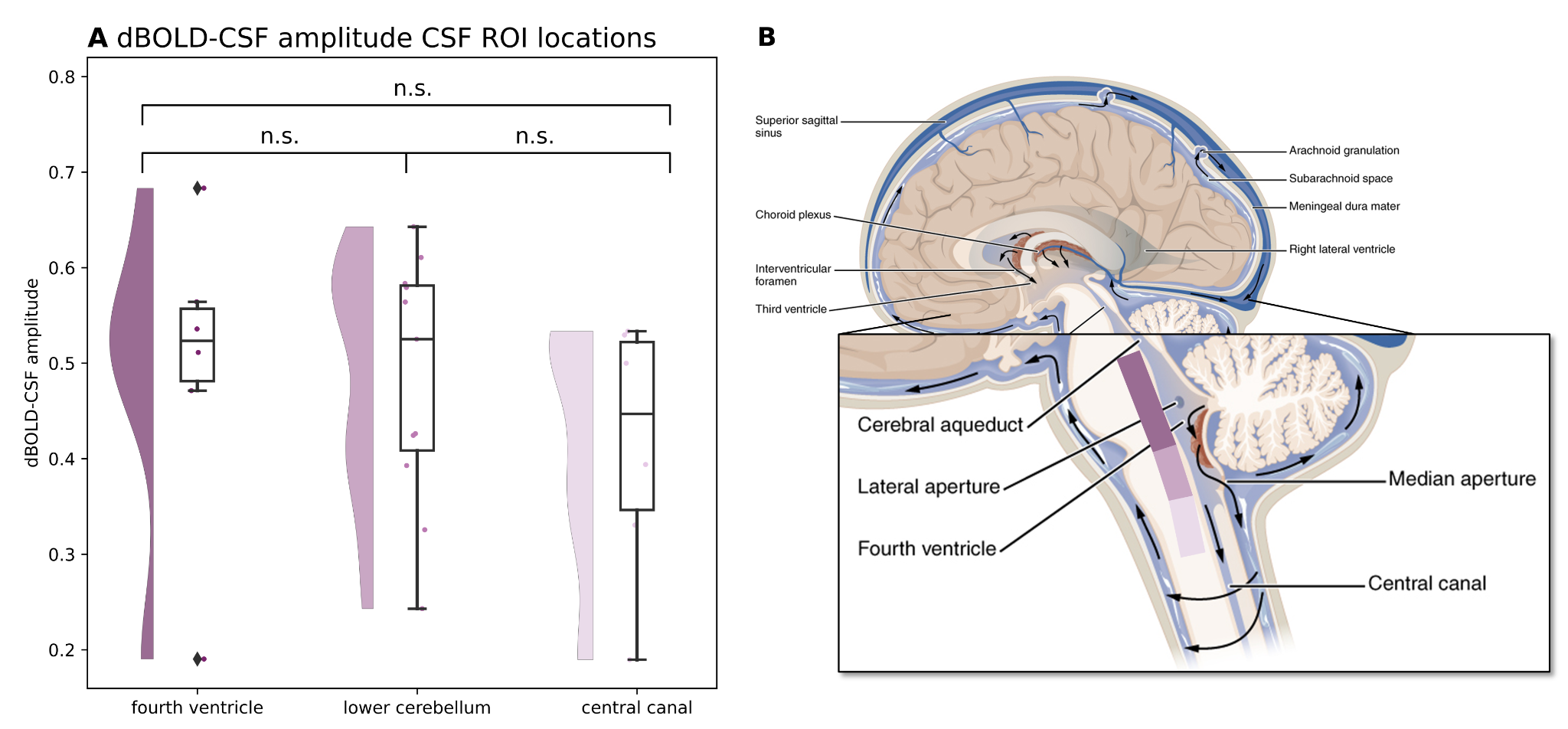

**SFig. 4. BOLD-CSF coupling amplitudes for different CSF ROI locations.** **(A)** Fourth ventricle-lower cerebellum *p*=.469; fourth ventricle-central canal *p*=.832; lower cerebellum-central canal *p*= .718. **(B)** Schematic representation of the anatomical locations: at the level of the fourth ventricle, lower cerebellum, and central canal. Different shades of purple correspond with the categories in panel A. Anatomical image is adapted from *OpenStax Anatomy and Physiology*, licensed under CC-BY 4.0, with modifications."
